# Urinary tract infection in infants with asymptomatic Jaundice: a meta-analysis UTI and Jaundice Review

**DOI:** 10.1101/2022.07.26.22278041

**Authors:** Anand Gourishankar, Smita. S. Akkinapally

**Affiliations:** Children’s National Hospital; Ballad Health

**Keywords:** Urinary tract infections, Systematic review, Meta-analysis

## Abstract

**INTRODUCTION:** Infants, especially neonates, present with jaundice with an unclear association with urinary tract infection. Evidence for such association is unclear, especially in a specific group of otherwise well-appearing infants born > 35 weeks.

**EVIDENCE ACQUISITION:** *Data sources:* We used the following databases: Medline, Embase, CINAHL Plus, Scopus, and Cochrane library.

*Study selection:* We included observational studies that included infants born > 35 weeks gestation, younger than 12 months, asymptomatic other than jaundice, and urinary tract infection. Data extraction: After reviewing the eligibility, two reviewers extracted data and assessed the quality of each study using the Newcastle-Ottawa scale.

**EVIDENCE SYNTHESIS:** We analyzed sixteen studies for a total of 2933 infants. The pooled incidence of UTI was 9.6% (95% confidence interval of 6% to 15%). The subgroup analysis failed to show any difference in incidence within the publication year, sample size, study design, study region, urine collection method, and age group. There was no explanation of heterogeneity noted by the meta-regression for UTI incidence with publication year, total bilirubin, sample size, and study quality. The funnel plot and Egger’s test revealed publication bias.

**CONCLUSIONS:** Nearly 1 in 10 otherwise asymptomatic infants with jaundice have a UTI. We recommend a rigorous large prospective study to confirm this finding.

## INTRODUCTION

The clinical significance of urinary tract infection (UTI) in infants presenting only with jaundice is uncertain. Jaundice appears in two-thirds of term newborns and almost 80% of preterm infants in the first weeks of life. The jaundice approach in infants, primarily neonates, depends on the age of onset, symptoms, type of hyperbilirubinemia, and risk factors.^1^ Sepsis, including urinary tract infection, is associated with jaundice. Concerns exist about the current practice of testing for UTI in otherwise asymptomatic infants presenting with jaundice. Expert committees (American Academy of Pediatrics-AAP, The National Institute for Health and Care Excellence-NICE, and Queensland Clinical Guidelines) have advised using clinical judgment when suspecting UTI in an asymptomatic infant with jaundice.^1-3^

Studies from the 1960s had described an association between jaundice and sepsis; since then, more studies are emerging.^4-6^ UTI is rare in newborns in the first three days of life.^7,8^ The overall incidence of UTI in infants ranged from 5% to 15%.^9,10^ Jaundice (up to 40%) is one of the clinical presentations in neonatal UTI. In general, clinical sepsis or neonatal fever, irrespective of jaundice, triggers a test for UTI. However, it is unclear whether to test for UTI in asymptomatic neonates and infants with only jaundice. Apart from indirect (unconjugated) hyperbilirubinemia, there is an association between direct (conjugated) hyperbilirubinemia and UTI.^11-13^ A 2018 systematic review reported that UTI prevalence in asymptomatic infants with jaundice was 7%, while other studies have reported between 0.6% and 53.9%.^14^ Recently there has been an increasing trend in the incidence of UTI in this group.^13,15-17^ The AAP guideline recommends testing for UTI in cases of direct hyperbilirubinemia based on a single study.^1,11^ However, other researchers have discouraged urine culture due to the low incidence of UTI.^18-21^

Current guidelines and evidence vary with the study region and the population. Given a lack of consensus regarding optimal practice, our objective was to determine UTI incidence in otherwise asymptomatic infants with jaundice born > 35 weeks gestation and under one year of age.

## EVIDENCE ACQUISITION

### Data sources, selection criteria, and search strategy

We registered our protocol with the international prospective register of systematic reviews (PROSPERO; registration number: CRD42016039129). The PRISMA guideline aided in performing the systematic review and meta-analysis.^22^ Two reviewers (A.G. and S.A.) screened titles and abstracts of the potentially relevant studies. The authors reconciled any disagreements of eligibility by discussions. The review included fifty-three full-text studies. We excluded 37 of them due to the wrong study design, missing information, and non-English language.

We included studies that reported the incidence of UTI in otherwise well-appearing infants presenting with clinical jaundice. Inclusion criteria were: (1) infants under 12 months and born after 35 weeks; (2) cohort and case-control studies; and (3) documented methods of urine collection. The rationale for age limit and gestation allowed us to compare publications before and after the American Academy of Pediatrics publication of the jaundice guideline.^1^ The authors excluded studies without information on sample size, UTI, gestational age, or methods used for diagnosing UTI. We did not include studies that reported clinical sepsis if the infant’s age was above 12 months or born before 35 weeks gestation. Our review excluded animal studies, studies published in a language other than English, and case reports.

A professional research librarian conducted the literature search. The librarian used MeSH headings (Jaundice, Neonatal Jaundice, Neonatal Hyperbilirubinemia, Urinary Tract Infections, Cystitis, Pyelonephritis, Pediatrics, and Infant) as well as equivalent keywords and phrases (direct and indirect bilirubinemia, conjugated and unconjugated bilirubinemia, physiologic jaundice, conjugated jaundice, direct and indirect jaundice, urinary bladder infections and kidney infections). The supplementary material (supplemental Figure 4) has the details of the search strategy. The librarian translated the search strategy from Medline Ovid to Embase (Elsevier), CINAHL Plus with Full Text (EBSCO), Scopus (Elsevier), and Cochrane Library (Wiley). The literature search ran until January 2020. There were a total of 1215 citations on the subject matter from all the databases. The librarian combined the citations into an EndNote Library 9.2 (Thomson Reuters), removed the duplicate studies, and exported them into Rayyan (Qatar Computing Research Institute) for the title, abstract, and full-text screening.^23^

### Data Extraction and quality assessment

After piloting a data extraction form, the two review authors agreed upon the final data extraction form. Two authors independently abstracted the data from studies, compared and resolved differences by discussion. We collected the following information in the data form: study identification (author, journal, country, study type, setting and year), study variables (period of study, age group, inclusions, exclusions, gestation-Term, preterm; <35wks/35-36wks, urine source - catheter /suprapubic/urine bag, UTI diagnostic method), number of infants with UTI, the total number of infants evaluated, and bilirubin (total and direct). We assumed ‘term’ as infants born more than 37 weeks unless specified. We used the Newcastle-Ottawa Scale (NOS) for quality assessment of observational studies (http://www.ohri.ca/programs/clinical_epidemiology/oxford.asp). This scale uses a star system to assess cohort and case-control studies separately (Table 2), and the maximum overall quality score possible is nine.

**Table I.**
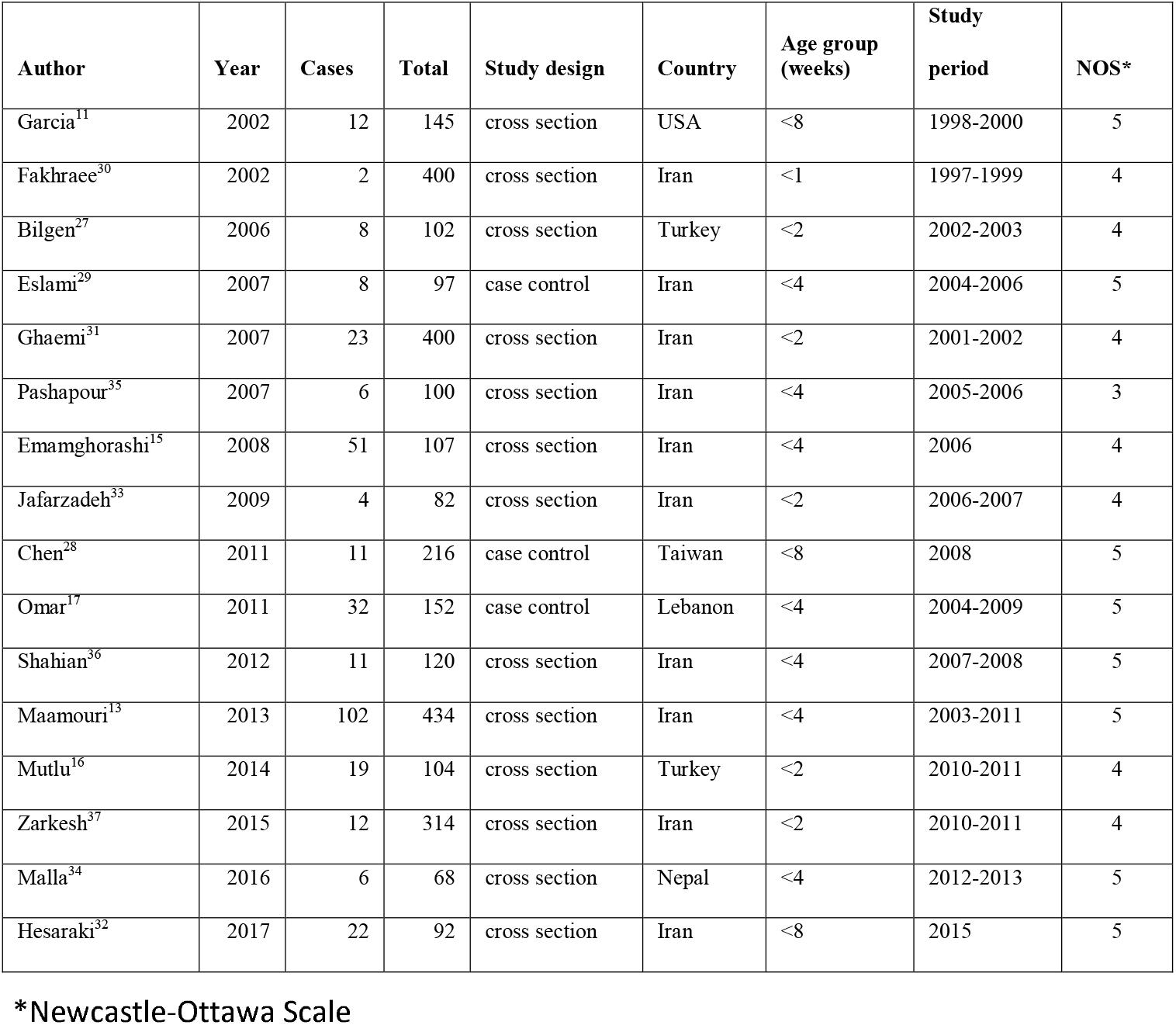
Characteristics of the included studies.

**Table II.**
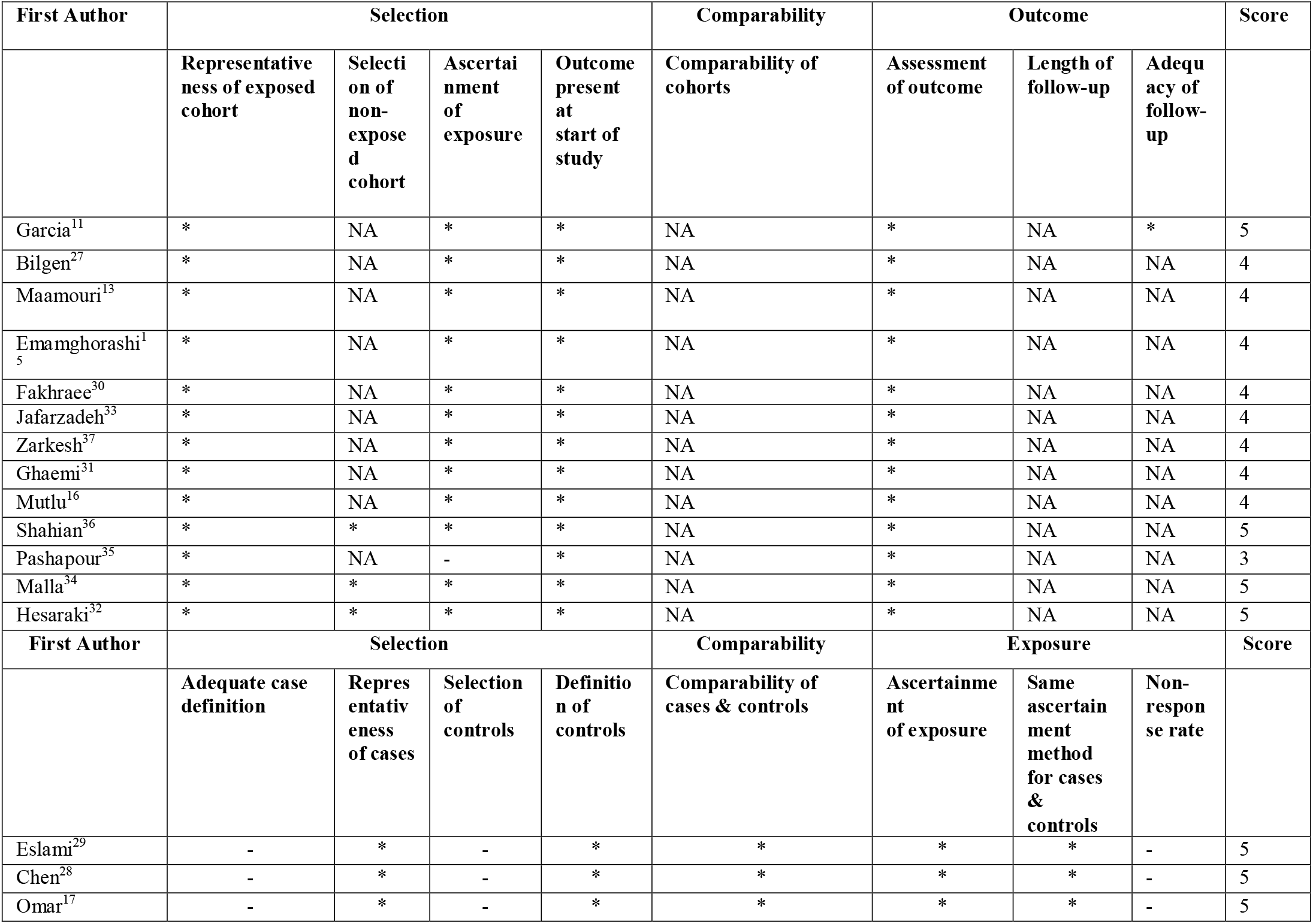
Quality assessment of the included studies based on the Newcastle-Ottawa scale

| First Author | Selection |  |  |  | Comparability | Outcome |  |  | Score |
| --- | --- | --- | --- | --- | --- | --- | --- | --- | --- |
|  | Representative<br>ness of exposed<br>cohort | Selecti<br>on of<br>non-<br>expose<br>d<br>cohort | Ascertain<br>ment<br>of<br>exposure | Outcome<br>present<br>at<br>start of<br>study | Comparability of<br>cohorts | Assessment<br>of outcome | Length of<br>follow-up | Adequ<br>acy of<br>follow-up |  |
| Garcia<sup>11</sup> | * | NA | * | * | NA | * | NA | * | 5 |
| Bilgen<sup>27</sup> | * | NA | * | * | NA | * | NA | NA | 4 |
| Maamouri<sup>13</sup> | * | NA | * | * | NA | * | NA | NA | 4 |
| Emamghorashi<sup>15</sup> | * | NA | * | * | NA | * | NA | NA | 4 |
| Fakhraee<sup>30</sup> | * | NA | * | * | NA | * | NA | NA | 4 |
| Jafarzadeh<sup>33</sup> | * | NA | * | * | NA | * | NA | NA | 4 |
| Zarkesh<sup>37</sup> | * | NA | * | * | NA | * | NA | NA | 4 |
| Ghaemi<sup>31</sup> | * | NA | * | * | NA | * | NA | NA | 4 |
| Mutlu<sup>16</sup> | * | NA | * | * | NA | * | NA | NA | 4 |
| Shahian<sup>36</sup> | * | * | * | * | NA | * | NA | NA | 5 |
| Pashapour<sup>35</sup> | * | NA | - | * | NA | * | NA | NA | 3 |
| Malla<sup>34</sup> | * | * | * | * | NA | * | NA | NA | 5 |
| Hesaraki<sup>32</sup> | * | * | * | * | NA | * | NA | NA | 5 |
| First Author | Selection |  |  |  | Comparability | Exposure |  |  | Score |
|  | Adequate case<br>definition | Representativ<br>eness<br>of cases | Selection<br>of<br>controls | Definitio<br>n of<br>controls | Comparability of<br>cases & controls | Ascertainme<br>nt<br>of exposure | Same<br>ascertain<br>ment<br>method<br>for cases<br>&<br>controls | Non-<br>respon<br>se rate |  |
| Eslami<sup>29</sup> | - | * | - | * | * | * | * | - | 5 |
| Chen<sup>28</sup> | - | * | - | * | * | * | * | - | 5 |
| Omar<sup>17</sup> | - | * | - | * | * | * | * | - | 5 |

| **First Author** | **Selection** |  |  |  | **Comparability** | **Outcome** |  |  | **Score** |
| --- | --- | --- | --- | --- | --- | --- | --- | --- | --- |
|  | **Representativeness of exposed cohort** | **Selection of non-exposed cohort** | **Ascertainment of exposure** | **Outcome present at start of study** | **Comparability of cohorts** | **Assessment of outcome** | **Length of follow-up** | **Adequacy of follow-up** |  |
| Garcia<sup>11</sup> | * | NA | * | * | NA | * | NA | * | 5 |
| Bilgen<sup>27</sup> | * | NA | * | * | NA | * | NA | NA | 4 |
| Maamouri<sup>13</sup> | * | NA | * | * | NA | * | NA | NA | 4 |
| Emamghorashi<sup>15</sup> | * | NA | * | * | NA | * | NA | NA | 4 |
| Fakhraee<sup>30</sup> | * | NA | * | * | NA | * | NA | NA | 4 |
| Jafarzadeh<sup>33</sup> | * | NA | * | * | NA | * | NA | NA | 4 |
| Zarkesh<sup>37</sup> | * | NA | * | * | NA | * | NA | NA | 4 |
| Ghaemi<sup>31</sup> | * | NA | * | * | NA | * | NA | NA | 4 |
| Mutlu<sup>16</sup> | * | NA | * | * | NA | * | NA | NA | 4 |
| Shahian<sup>36</sup> | * | * | * | * | NA | * | NA | NA | 5 |
| Pashapour<sup>35</sup> | * | NA | - | * | NA | * | NA | NA | 3 |
| Malla<sup>34</sup> | * | * | * | * | NA | * | NA | NA | 5 |
| Hesaraki<sup>32</sup> | * | * | * | * | NA | * | NA | NA | 5 |
| **First Author** | **Selection** |  |  |  | **Comparability** | **Exposure** |  |  | **Score** |
|  | **Adequate case definition** | **Representativeness of cases** | **Selection of controls** | **Definition of controls** | **Comparability of cases & controls** | **Ascertainment of exposure** | **Same ascertainment method for cases & controls** | **Non-response rate** |  |
| Eslami<sup>29</sup> | - | * | - | * | * | * | * | - | 5 |
| Chen<sup>28</sup> | - | * | - | * | * | * | * | - | 5 |
| Omar<sup>17</sup> | - | * | - | * | * | * | * | - | 5 |

### Data synthesis and analysis

The pooled incidence was estimated and displayed in the forest plots with 95% confidence intervals. We used inverse variance when calculating the weights of the individual studies. The logit transformation of proportions produced a normal distribution (as proportions are less than 0.2).^24^ The random-effects model, which takes both within- and between-study variances into account, was applied using the DerSimonian and Laird (DL) method. We expected considerable heterogeneity due to observational studies.^25^ Heterogeneity was categorized as follows: not important (0% - 40%); moderate (30% - 60%), substantial (50% - 90%); and considerable (75% - 100%).^26^ Publication bias was assessed visually by funnel plot and statistically by Egger’s test. Subgroup analysis included world region, research period (<2004/>2004; AAP guidelines published in 2004), age groups, study design, and urine collection method. Author, AG, conducted the meta-analysis using R software 3.2 (The R Foundation). Meta-regression analyzed the following variables: year published, country of publication, quality of the study, sample size, and total bilirubin and assessed for statistical significance (p ≤ 0.05). We performed a sensitivity analysis using the Leave-One-Out function, which graphically shows outliers as the farthest displacement of each study’s summary proportion. Leave-One-Out analysis technic recalculates the results of meta-analysis each time, leaving out one study. We compared the pooled incidence of UTI with and without outliers identified in the sensitivity analysis. Our review did not require ethics approval, as this is a secondary analysis of published data.

## EVIDENCE SYNTHESIS

A total of 16 studies were eligible for inclusion, confirmed by the two authors (Figure 1). The total sample size was 2933 infants; Table 1 depicts the study characteristics. ^11,13,16,17,27-37^ The median sample size was 114 (IQR 99 - 241; range: 68 - 434). Out of 16 studies, thirteen were cross-sectional studies (81%), and three were case-control studies (19%). All studies had infants admitted to the hospital (neonatal unit) irrespective of enrollment either in the outpatient or emergency department. Publications originated from the following countries; Iran (10, 62%), Turkey (2, 12%), and one from the U.S.A., Lebanon, Nepal, and Taiwan. Although the inclusion age criterion was up to 12 months, studies with various age groups enrolled were < 2 weeks (6,37%), < 4 weeks (7, 44%), and < 8 weeks (3, 19%). Studies published before AAP guidelines (before 2004) were 4 (25%) compared to those studies after the year 2004 (12, 75%). The mean for the NOS quality score was 4.4 (SD 0.6). This mean quality score is relatively low, given the overall maximum score of nine. Among the included studies, Garcia et al.’s quality-score was high (5) because infants had follow-up after discharge. On the other hand, Shahian et al. had additional information on infants without jaundice diagnosed with UTI (non-exposed). Pashpouri et al. scored the lowest quality score (3) because of the unclear ascertainment of exposure (Table 2).

**Figure 1:**
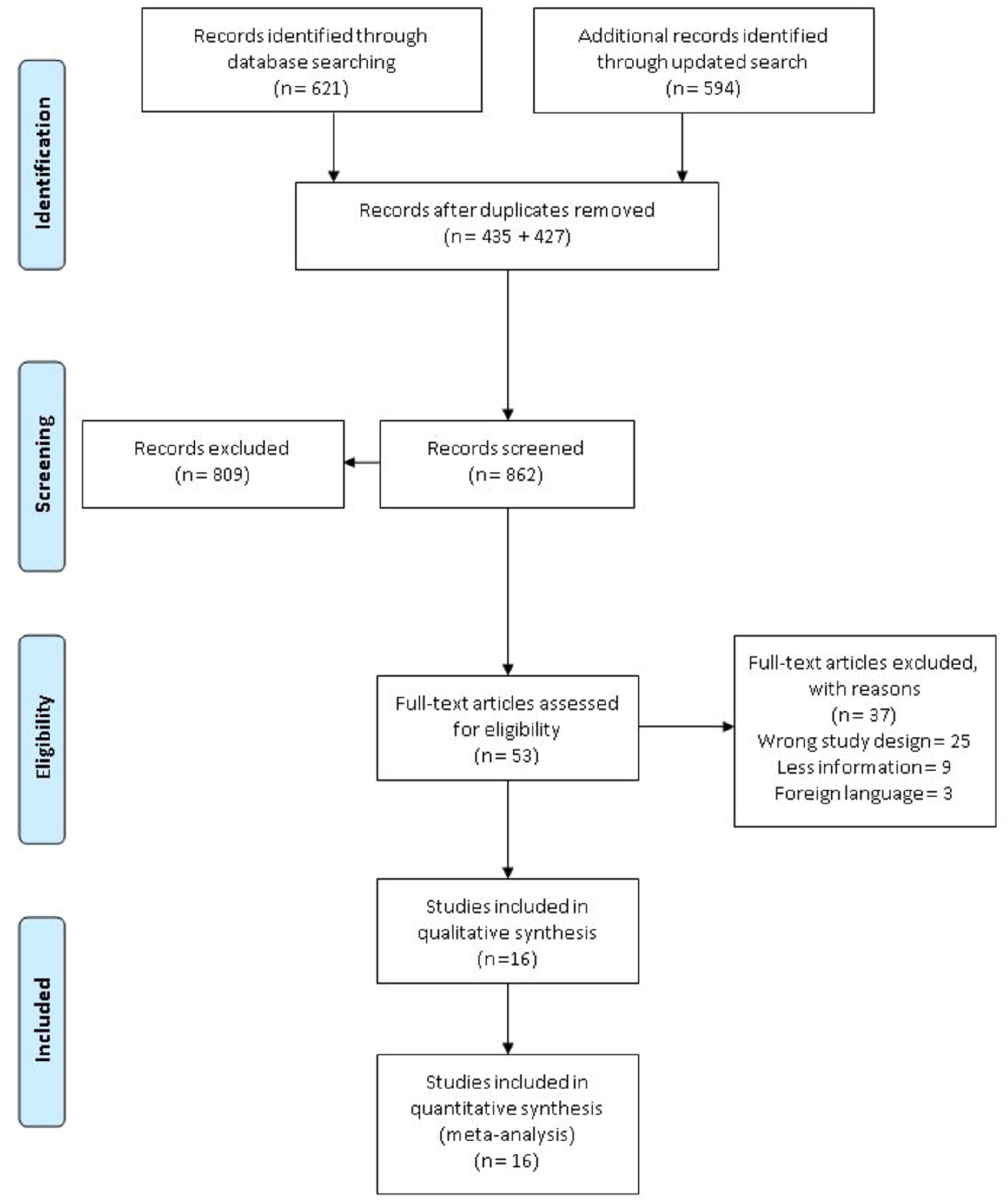
PRISMA flow diagram of the study selection.

UTI’s average raw incidence in infants (up to 8 weeks) with asymptomatic jaundice was 12.7%. The pooled incidence rate of UTI in an otherwise asymptomatic infant presenting with jaundice was 9.6% (95% CI: 6 to 15%; Figure 2). The heterogeneity was substantial (I^2^ = 93%).

**Figure 2:**
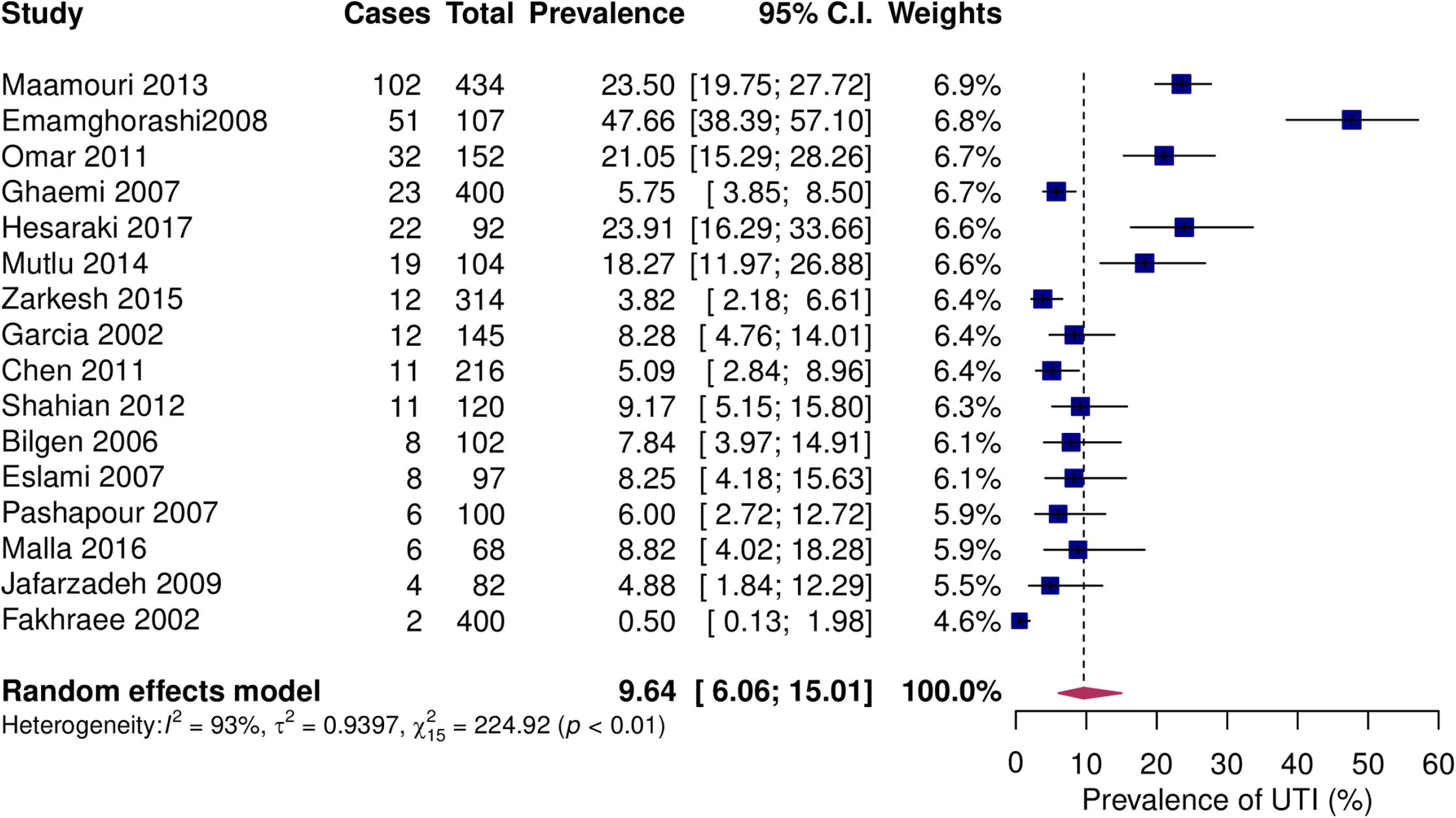
Forest plot of studies showing the prevalence of UTI in asymptomatic jaundiced infants. Ten studies display their point estimates, and the confidence interval overlaps with the pooled estimate and confidence interval.

The funnel plot (Figure 3) showed asymmetry when assessed visually, which may arise from heterogeneity, publication bias, or another factor.^38^ The right image in Figure 3 was evident for small-study-effect in this meta-analysis. Based on the sample size, smaller studies appeared around the vertical bar than the larger studies. Egger’s regression test confirmed that the funnel plot was significantly asymmetrical (z = -4.026, p < 0.001), indicating publication bias contributing to the low quality of evidence.

**Figure 3:**
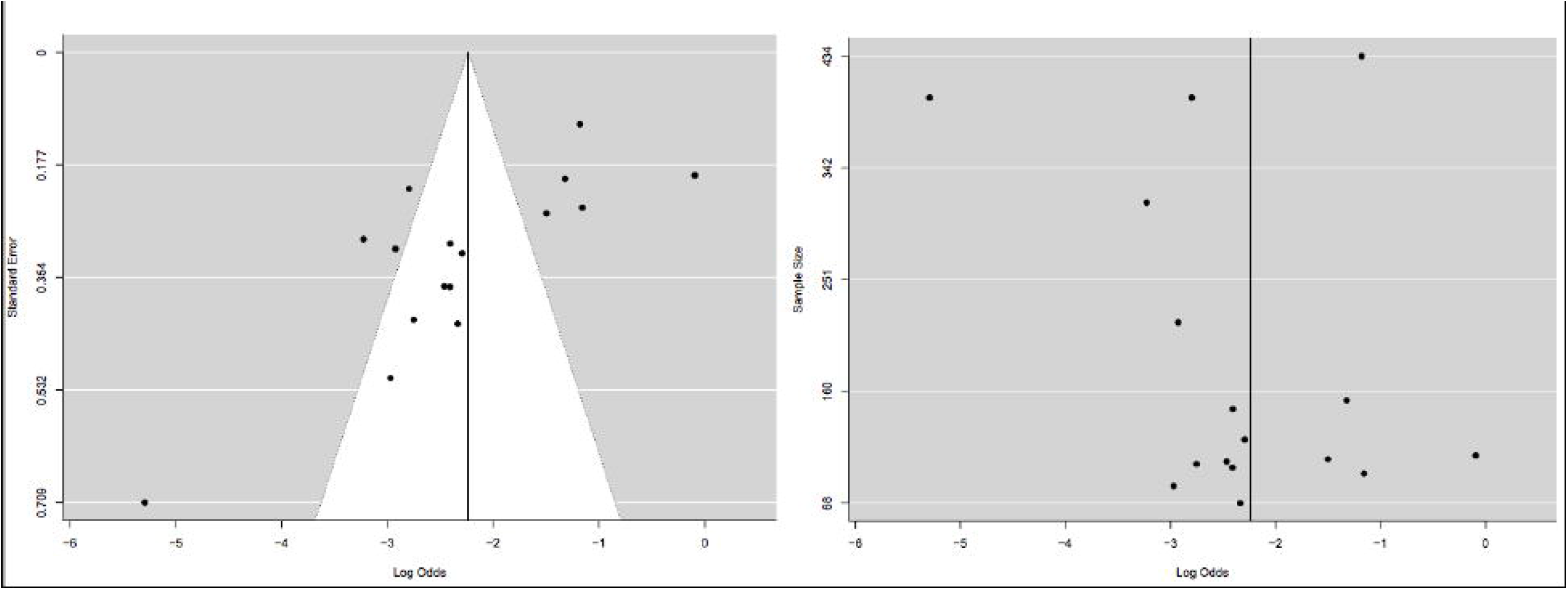
Funnel plot and scatter plot. Diagnostic funnel plot for publication bias. Each point represents a separate study. Asymmetric point distribution indicates the existence of publication bias. Scatter plot using sample size as the measure of precision to show the small-study effect. Points of asymmetry of large studies on one side.

Subgroup analyses for UTI incidence among the world regions, research period, age, study design, and urine collection method are shown in supplement, figure 4. These five subgroup analyses demonstrated variation in incidence between subgroups, where each analysis had considerable heterogeneity.

### Sensitivity Analysis

A sensitivity analysis of 16 studies identified two studies^15,30^ as an outlier (supplement, figure 2). The pooled incidence comparing with and without these two outliers was similar (9.6% (95% CI: 6 to 15%) versus 10.5%, 95% CI: 6.5 to 16.5%), with an overlap of confidence intervals.

### Meta-regression

After visual inspection of the Forest plot and subgroup analysis of the included studies, we examined five covariates (year published, country of publication, quality of the study, sample size, and total bilirubin) for potential heterogeneity. Meta-regression revealed no significant association between these variables and UTI incidence (supplement, Table 3).

## Discussion

We performed a systematic review and meta-analysis to investigate the incidence of urinary tract infection in asymptomatic infants with jaundice born after 35 weeks of gestation. Our review of 2933 infants from 16 studies showed a UTI pooled incidence of 9.6%. Our review’s UTI incidence was higher than the infants with asymptomatic bacteriuria. In comparison, a meta-analysis of fourteen studies done between the 1960s and 1980s found that the incidence of asymptomatic bacteriuria was 0.31% (95% CI; 0.21 % to 0.44%).^39^ Of note, our review’s UTI incidence was comparable to those with febrile UTI. The pooled incidence of febrile UTI among infants aged 0 to 3 months were as follows: females, 7.5%; circumcised males, 2.4%; and uncircumcised males, 20.1%.^10^ The weight of evidence suggests an association between UTI and otherwise asymptomatic infants presenting with jaundice. Based on the current evidence, we find the relevance of testing urine culture in otherwise well-appearing infants with non-hemolytic jaundice undergoing phototherapy until further research.

Subgroup analysis showed that a single nation had a lower incidence than other nations (9% vs. 11%), but meta-regression results were not significant. A systematic review published a similar result (7%, 95% CI; 5 to 11%).^14^ This systematic review was limited to prolonged jaundice, a single nation, included preterm and term, and the heterogeneity was considerable (I^2^ = 94%). By contrast, our review included multiple nations, only infants born > 35 weeks and age up to 8 weeks. The lower incidence (0.2%) in Steadman’s meta-analysis can be explained by the use of a fixed-effects model with the unclear transformation (with zero proportions), included letters and conference abstracts, and it had a substantial heterogeneity (I^2^ = 89.4%).^21^ Our review had rigorous methods, practical inclusion criteria (infant born > 35weeks, asymptomatic), and updated studies compared to the above two reviews.

Subgroup analysis by age showed divergent results. Thirty percent of studies included infants < 2 weeks, which had the lowest incidence compared to those enrolled < 4 weeks (44%) or < 8 weeks (19%). A lower incidence in the “< 2 weeks” group compared to “< 4 weeks” can be partly explained by the outliers identified in this study.^15,30^ Previous studies have described asymptomatic infants with jaundice and UTI in newborns after eight days.^7^ However, Omar et. al and Shahian et al. described contradictory findings (onset of jaundice < 7days with UTI).^17,36^ Therefore, we recommend testing for UTI in infants presenting with jaundice at ages from 3 days to 8 weeks. A higher UTI incidence after AAP jaundice guideline publication may reflect improved knowledge and result in more testing despite our non-significant meta-regression findings based on the publication year (supplemental Table 1).

UTI incidence was lower in the “invasive” group (catheter or suprapubic aspirate) than others. We must be cautious in interpreting the UTI incidence because sterile urine collection is essential. A more extensive prospective study with sterile urine collection and the definition of UTI would strengthen future meta-analysis.

### Strengths and Limitations

Limitations in our study stem from biases that can overestimate the incidence of UTI in asymptomatic jaundiced infants. There is no optimal definition of UTI diagnosis in neonates, and the AAP UTI guideline is limited to the age group that ranges from 2 months to 24 months.^40^ A substantial heterogeneity might arise from the clinical or methodological differences, other than chance. Clinical jaundice, as noticed by parents or providers, is subject to variability. Overall, the methodological quality of the included studies was low. Variation can occur in the study designs (inclusion criteria such as age, testing for other etiologies of jaundice), measurements (urine collection methods, laboratory standards, the timing of bilirubin testing), and demographics (ethnicity, gender, circumcision status). Most studies eliminated other jaundice causes, such as hemolysis (ABO, Rhesus, G6PD) and hypothyroidism, which helped support UTI and jaundice association. Therefore, we adopted a random-effects model due to heterogeneity, variability in the urine collection method, and diagnostic criteria. The other limitation was no access to individual patient data. We could not ascertain UTI diagnosis, fever status, and clinical jaundice. Our review was limited to the English language, and we did not include case series, poster presentation, or conference proceedings. Despite the limitations, this unique review included global studies, asymptomatic infants born after 35weeks, and rigorous methodological systematic review and meta-analysis, including adhering to the PRISMA guidelines, comprehensive search, and independent assessment.

## CONCLUSIONS

Urinary tract infection are associated with jaundice in otherwise well infant. Our review findings indicating a higher percentage of incidence raises concern and underscores the need for rigorous prospective studies examining the association between UTI and jaundice in infants.

## Data Availability

All data produced in the present study are available upon reasonable request to the authors.

## Authors’ contribution

Dr. Gourishankar made substantial contributions to the conception, acquisition, analysis, and interpretation of the data for the work. He also drafted the manuscript and revised it critically for important intellectual content.

Dr. Akkinapally made substantial contributions to the conception, acquisition, and interpretation of the data for the work. She also drafted the manuscript and revised it critically for important intellectual content.

Both authors read and approved the final version of the manuscript.

## Acknowledgements

We acknowledge methodological input from Mohan Pammi M.D., Ph.D. for the review. We thank Ms. Beatriz Varman, BS, MLIS, for her contribution in reviewing search methods, writing literature search methods, conducting a literature search, and consolidating studies into the software.

