## Supplementary material for "Urinary tract infection in infants with asymptomatic Jaundice: a meta-analysis UTI and Jaundice Review": suppl_Table3_metaregression.docx

**Table 3. Meta regression analysis of variables of heterogeneity in prevalence of UTI in asymptomatic infants with jaundice**

Variable	Univariate model

Coefficient	SE	p – value

Year of study	0.0956	0.0593	0.106

Country	-0.1595	0.5501	0.772

Quality (NOS)	0.4461	0.4388	0.309

Sample size	-0.0029	0.0023	0.204

Bilirubin	0.0042	0.0850	0.960
