## Supplementary material for "Urinary tract infection in infants with asymptomatic Jaundice: a meta-analysis UTI and Jaundice Review": Supplement_fig4_fig5_table4.docx

Supplemental Table 4: Medline Ovid Search Strategy

1. exp Jaundice/

2. exp Jaundice, Neonatal/

3. exp Hyperbilirubinemia, neonatal/

4. Hyperbilirubinemia/

5. (hyperbilirubinemia or bilirubinemia or indirect bilirubinemia or direct bilirubinemia or conjugated bilirubinemia or unconjugated bilirubinemia).mp.

6. (jaundice or physiologic jaundice or direct jaundice or indirect jaundice or conjugated jaundice or unconjugated jaundice).mp.

7. exp Urinary Tract Infections/

8. (uti or urinary tract infection$1 or bladder infection$1 or urinary bladder infection$1 or cystitis or pyelonephritis or acute pyelonephritis or kidney infection$1).mp.

9. exp Cystitis/

10. exp Pyelonephritis/

11. 1 or 4 or 5 or 6

12. 7 or 8 or 9 or 10

13. 11 and 12

14. exp pediatrics/ or exp infant/ or (infancy or pediatric$1 or infantile or neonatology or infant$ or bab$3 or newborn$1 or neonate$1 or neonatal or infant newborn).mp.

15. 13 and 14

16. (2 or 3) and 12

17. 15 or 16

18. (neonatal jaundice or neonatal hyperbilirubinemia).mp.

19. 18 and 12

20. 17 or 19

**Figure 4a: Forest plot for studies published in Iran (nation =1) and**

**other nations (nation =0)**

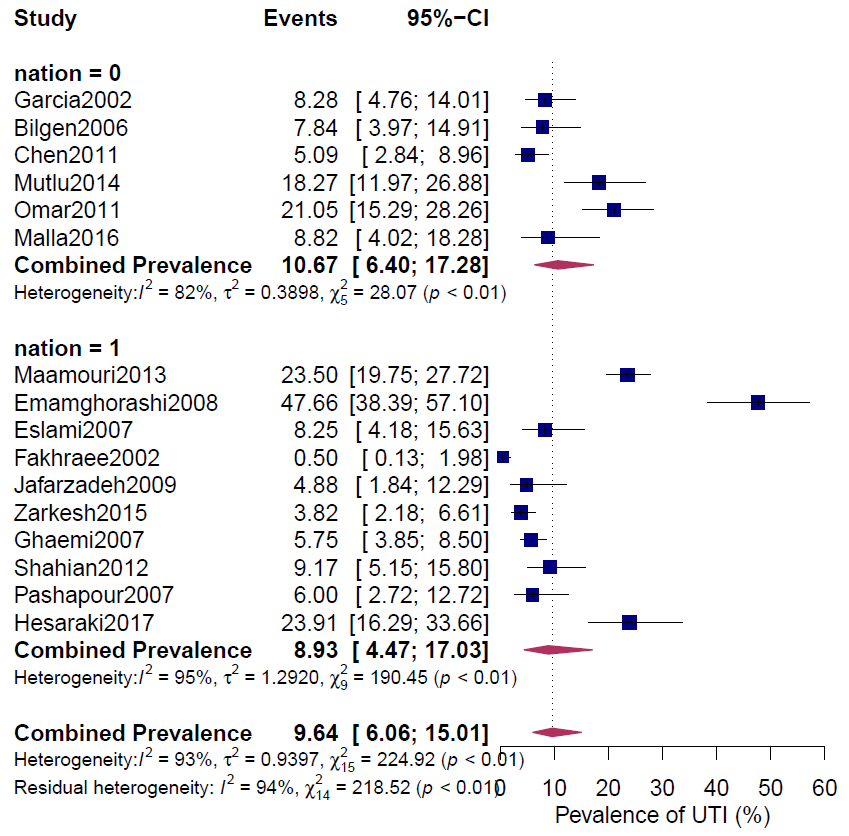

**Figure 4b: Forest plot for the comparison of studies before and after AAP jaundice guideline publication**

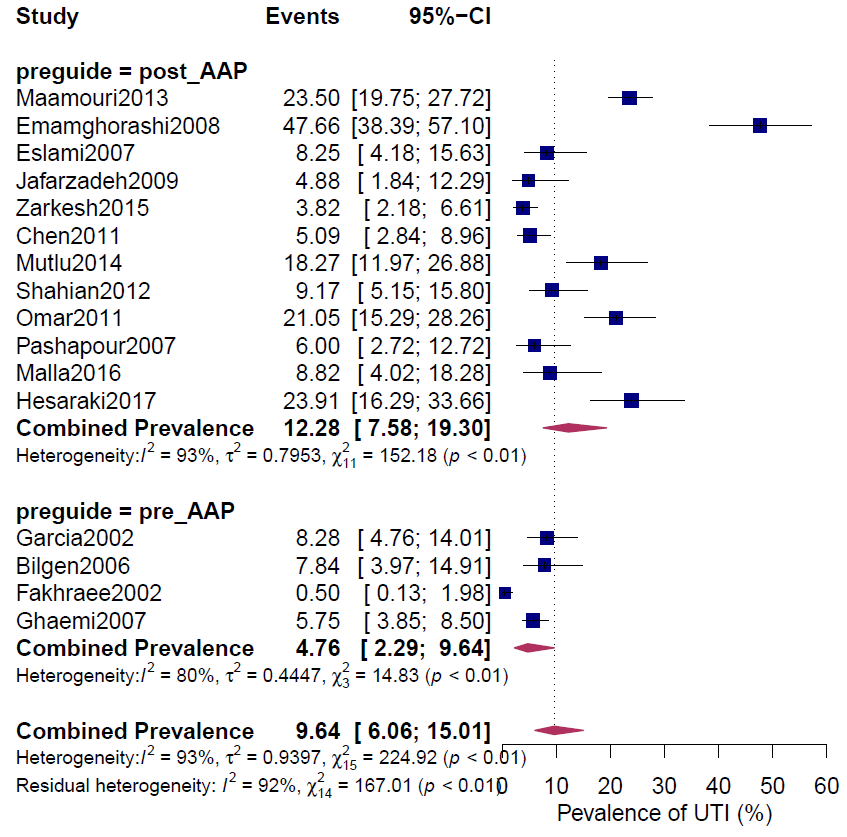

**Figure 4c: Forest plot comparing studies grouped by age**

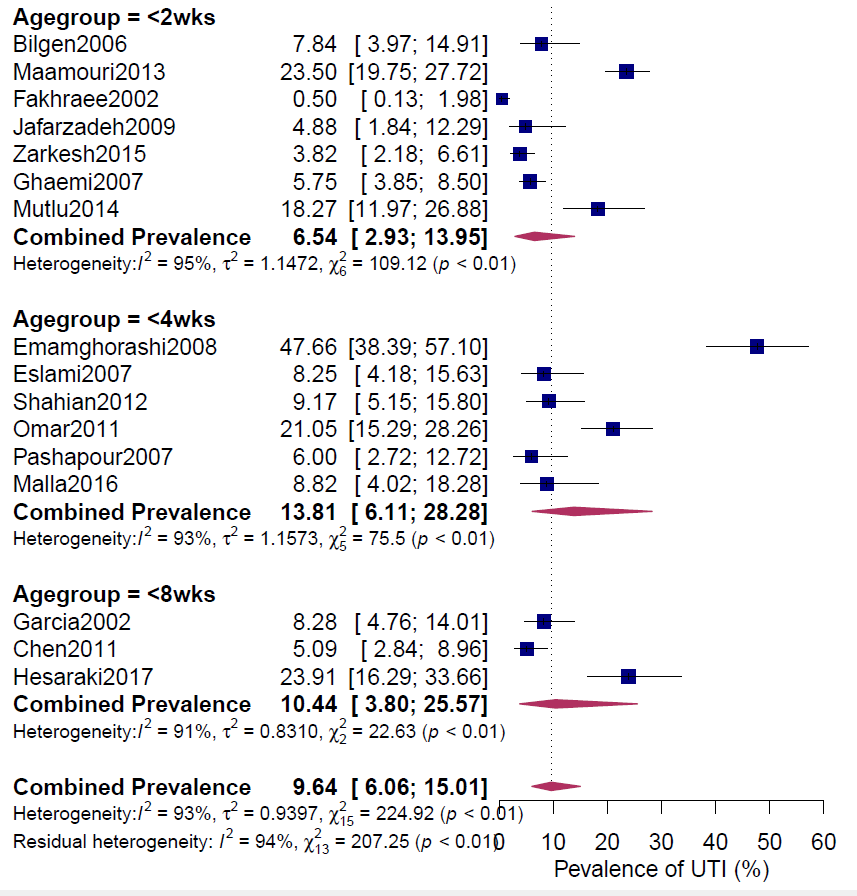

**Figure 4d: Forest plot comparing the type of study designs**

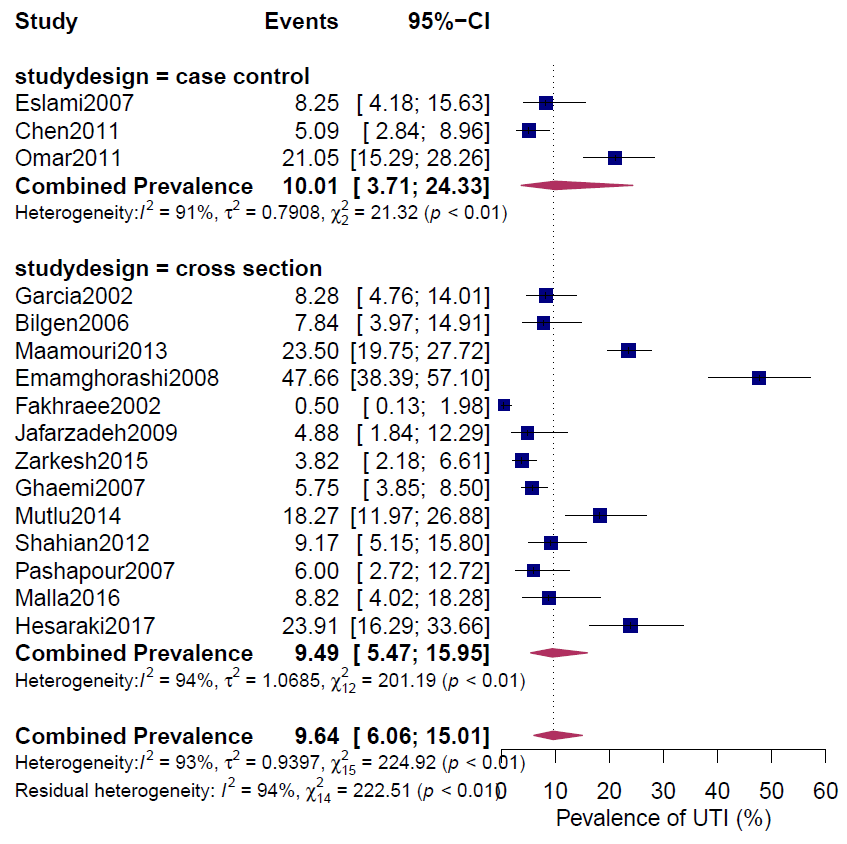

**Figure 4e: Forest plot for comparing studies based on urine collection methods**

**(invasive = SPA/Catheter, other = Bag/SPA/Catheter).**

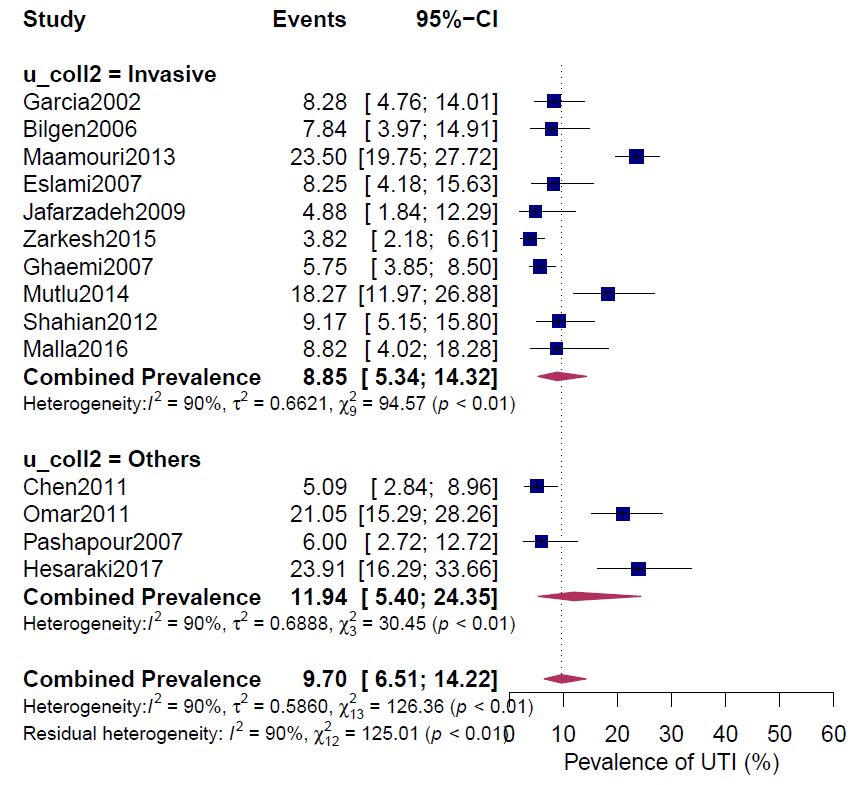

**Figure 5: Sensitivity Analysis. Effect of each study as an outlier (left out) on the proportion (outliers are farther from midline)**

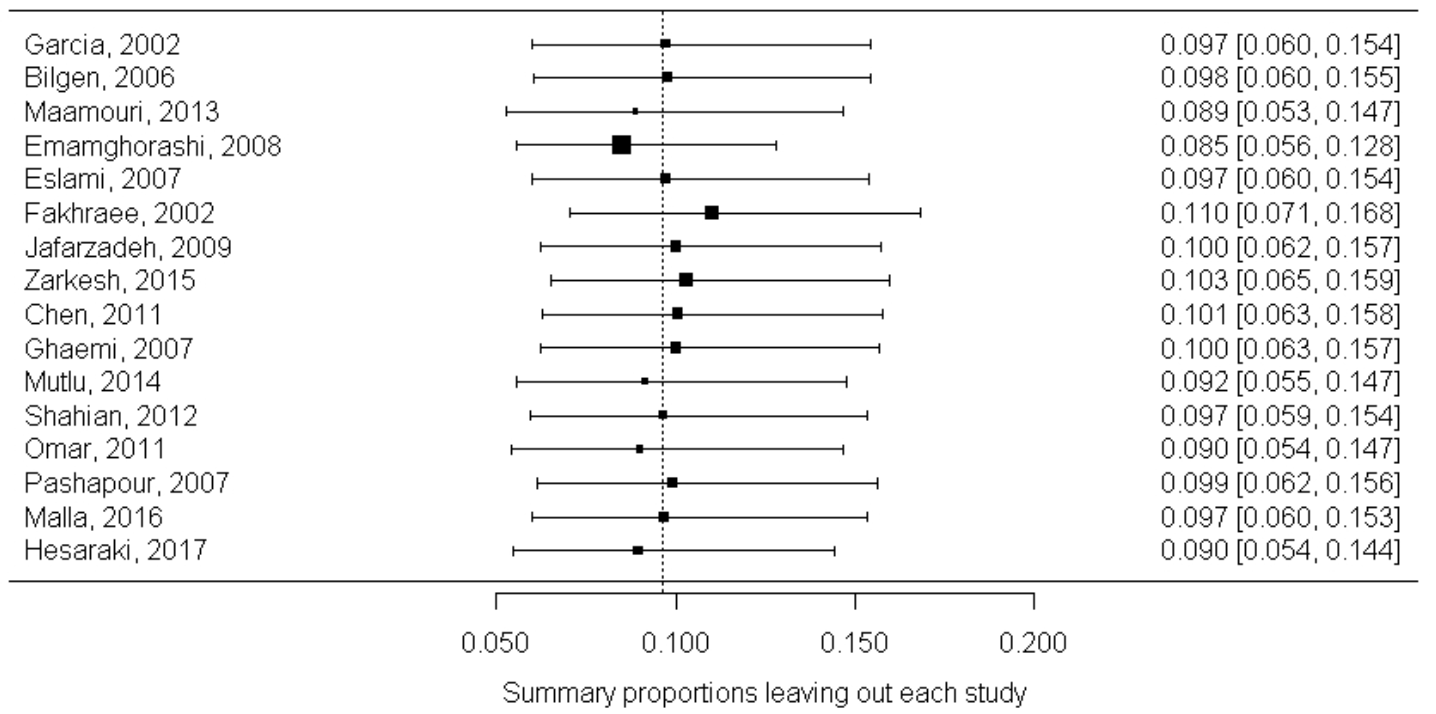
